# Impact of health systems interventions in primary health settings on type 2 diabetes care and health outcomes among adults in West Africa: a systematic review protocol

**DOI:** 10.1101/2023.08.31.23294889

**Authors:** Eugene Paa Kofi Bondzie, Yasmin Jahan, Dina Balabanova, Tony Danso-Appiah, Tolib Mirzoev, Edward Antwi, Irene Ayepong

## Abstract

Type 2 diabetes remains a major global public health challenge particularly in the African region. Though evidence exists on pharmacological agents and non-pharmacological interventions in maintaining blood glucose concentration, the health systems ability in meeting patients’ needs may be inadequate. However, the management of non-communicable diseases particularly diabetes, have been postulated to depend largely on functioning health systems. This systematic review will therefore, summarize the current evidence on existing health systems interventions in primary health settings for type 2 diabetes care and health outcomes in West Africa and would explore the impact of these system-level interventions on service availability, accessibility and quality, as well as individualized outcomes such as glycemic control, disease awareness and treatment adherence.

The review will be conducted in accordance with the reporting guidance provided in the Preferred Reporting Items for Systematic Reviews and Meta-Analyses Protocols (PRISMA-P). The health system framework by Witter et al, 2019 will guide the system-level interventions and the search strategy to be explored in this review. This framework was designed to integrate the six building blocks of the World health’s organization (WHO) health systems framework and delineates how they work synergistically to improve specific health outcomes. We will search the following databases PubMed, Google scholar and Cumulated Index to Nursing and Allied Health Literature (CINAHL) between January 2000 to June 2023 and Car.info from inception to June 2023. The Cochrane Collaboration tool for assessing Risk of Bias will be implemented on each included study to assess for risk of bias.

We will conduct a narrative synthesis and make comparisons across findings using Excel generated tables.

The main limitation of this study is that we are likely to miss out on studies not conducted in English or French since our search would be conducted in English and French only.

**In Conclusion,** this systematic review will outline the existing system-level interventions that aim to or already improve type 2 diabetes services in primary health facilities in West Africa and would allow for strengthening and co-production of successful interventions that can be generalized to the entire sub-region.

## Introduction

Type 2 diabetes, formerly known as adult-onset diabetes, is a major global public health problem(1). It is defined as a form of diabetes mellitus that is characterized by high blood glucose, insulin resistance and a relative lack of insulin and it manifests with symptoms of increased thirst, frequent urination, weight loss and sometimes increased hunger(2). Long term complications from high blood sugar includes ischemic heart disease, retinopathy, nephropathy and limb amputations(3).

Maintaining blood glucose concentration is highly essential to prevent severe complications(4). Until recently, it was thought to affect adults who were middle-aged or older but recent trends have shown an increase incidence in young people(5). Reports from epidemiological studies suggest that adults aged 20-79 years living with type 2 diabetes are about 10.5% of the world’s population(6). In Sub-Saharan Africa, the disease is estimated to increase by 129% by 2045 due to an increase in population and rapid urbanization(6). This phenomenon is worsened by the large number of undiagnosed individuals living with the disease. According to published reports from the international diabetes federation (IDF) Atlas, people with undiagnosed diabetes in the African region represent the highest comparative proportions worldwide, currently at 54% and expected to increase by 2045(6). Consequently, the number of diabetes related death was reported at 416,000 compared with 111,100 deaths in Europe. These disproportionate number reflect the poor structures at various levels of the primary care management of non-communicable diseases in the African region(7,8). Evidence exists on the effectiveness of pharmacological and non-pharmacological treatment for type 2 diabetes(9,10,11) but the health systems ability in meeting patients’ needs may be inadequate(2,7).

The management and control of type 2 diabetes is largely dependent on health systems(12). The treatment of diabetes is challenging because health systems are designed for short-term care rather than long-term care of people(13). A healthcare system is an organization of people, institutions, and resources that delivers health care services to meet the health needs of target populations(14).

Effective health systems can improve the delivery of diabetes care and promote patient’s access and utilization of quality services including access to medications, health facilities and specialists, with an overall impact on glycemic control and other associated outcomes (e.g., adherence, morbidity and mortality etc.)(14,15,16).

A previous systematic review was conducted by Nuche-Berenguer, et al (17)on the readiness of health systems in tackling diabetes in Sub-Saharan Africa, with part of the review focused on evaluating pilot projects, targeted at enhancing the capacity of health systems in diabetes care. However, this review was conducted six years ago, at a time when health system interventions towards type 2 diabetes were now evolving in most Sub-Saharan African countries especially West-African countries. Thus, the majority of the studies identified in that review were based-off eastern and southern Africa and the pooled findings may not be necessarily generalizable in recent times to other parts of Africa. The review also could not synthesize the impact of the interventions on glycemic control due to the scarcity of relevant papers.

However, studies have been conducted between that period and the present to determine the impact of health system strengthening interventions on glycemic control. These include strategies aimed at improving the training and distribution of healthcare personnel, improving patient follow-up appointments, improving medicines distribution, and decentralizing services to patients at the primary-care level etc. Also, diversity exists between the sub-populations, their economic growth, health facility distribution, general disease burden and political structures. Thus, it will be important to determine which interventions have been practical in the West African sub population.

According to the World Health Organization (WHO), health systems interventions can be defined as any array of initiatives that improves one or more of the functions of the health systems and that leads to better health through improvements in access, coverage, quality or efficiency(18). Chee et al, proposed that these initiatives should aim at permanently making the systems function better, not just filling gaps or supporting the systems to produce better short-term outcomes. They further submit that an intervention to strengthen the health system should go beyond providing inputs and apply to more than one building block, citing the six core components of health systems identified by the WHO.

For the purposes of this review, the health system interventions framework by Witter et al(18) in 2019 would guide the system-level interventions to be explored in this review (S1 Fig). This framework was designed to integrate the six building blocks of the WHO health systems framework (14). The framework by Witter et al, describes mechanisms of change within six health system blocks (governance, financing, infrastructure, workforce, supply chain and information), describes the implementation process goals and outlines the final desired outcomes. Interventions that promote any of these domains of the health system or across integration of the domains, have been proven to have significant impact on the outcome of a health condition (non-communicable disease) in the African setting(19).

This systematic review will summarize the recent evidence on health system interventions implemented in primary care facilities to improve the accessibility, delivery and quality of healthcare for type 2 diabetes in West Africa; and will explore the impact and effectiveness of these interventions on overall glycemic control, disease awareness and treatment adherence.

**S1 Fig.**
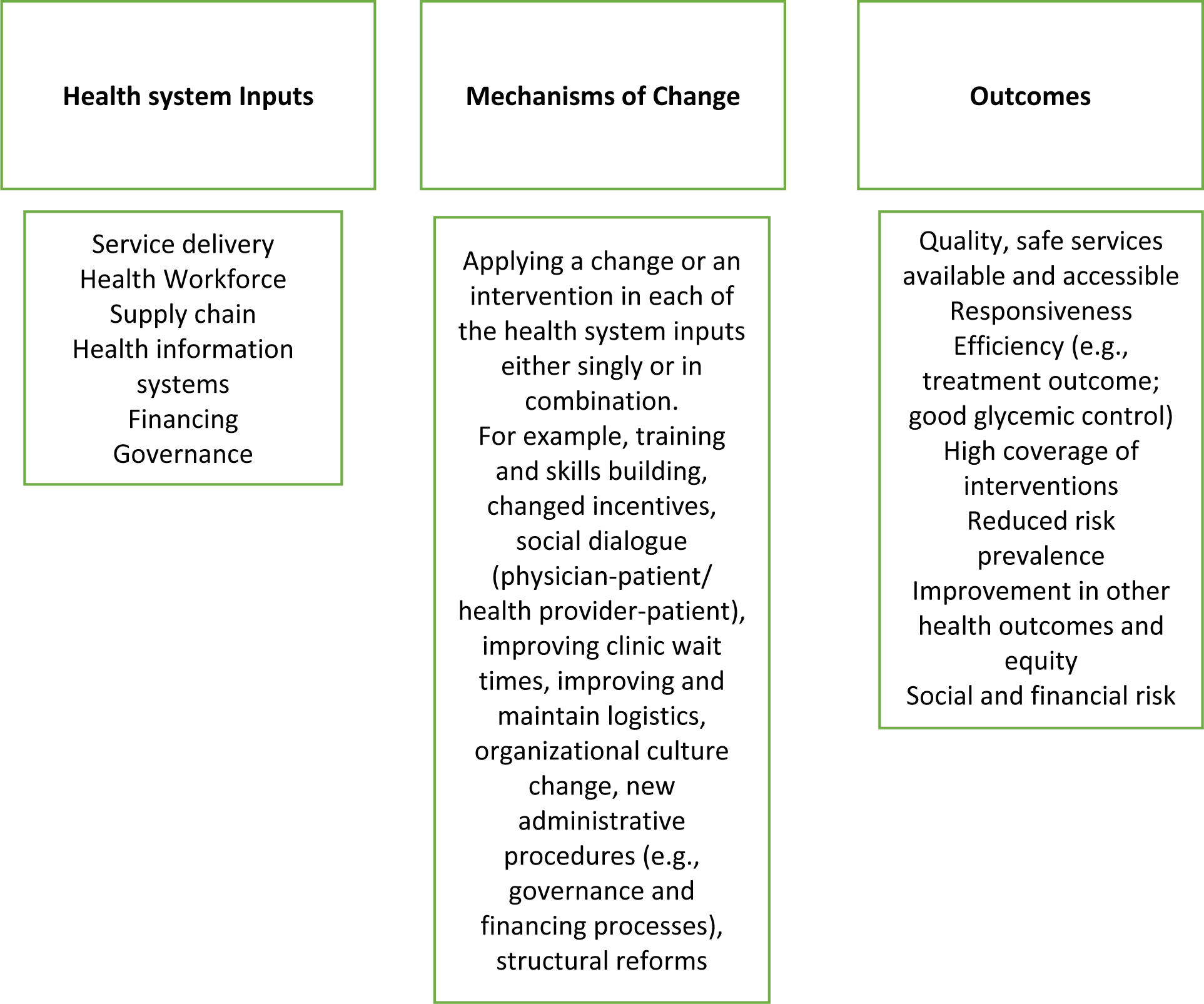
Health systems interventions framework (Modified from the health systems strengthening framework by Witter et al(18))

### Review question

This systematic review aims to answer the question;

How do health systems interventions influence care and health outcomes of type 2 diabetes in primary health settings among adults in West Africa?

### Objectives

#### General

To identify the health system interventions in primary health settings that influence the accessibility, delivery and quality of type 2 diabetes care among adults in West Africa.

#### Specific

1. To synthesize the evidence on the effectiveness of these system-level interventions on glycemic control, awareness and treatment adherence.

2. To explore the impact of these system-level interventions on any associated health outcomes (e.g., service availability, service utilization etc.)

## Methods

This systematic review will follow the reporting guidance provided in the Preferred Reporting Items for Systematic Reviews and Meta-Analyses Protocols (PRISMA-P) statement (20) (S1 Checklist)

### Criteria for considering studies for this review

#### PICOS eligibility (S1 Table)

##### Types of studies

- We will include studies with randomised controlled trials (RCTs) trials, clinical control trials (CCTs) and quasi-experimental designs
- Observational studies including cross-sectional studies, cohort studies, case-control studies and case series will be excluded
- Gray literature (e.g., books, commentary, dissertations, conference proceedings, modeling and simulation studies) will also be excluded.

##### Types of participants and setting

- The participants to be covered in this review will be adults aged 18 years or more in West Africa with a diagnosis of type 2 diabetes.
- Studies conducted in a primary health facility setting (community health and planning services (CHPS) compound, health center, district hospital/clinic) will be eligible
- Studies on gestational diabetes (since its natural history varies significantly from type 2 diabetes(21)), and type 1 diabetes will be excluded
- Also, studies that evaluated both type 1 diabetes and type 2 diabetes together will be excluded, if we cannot extract findings from the group with type 2 diabetes.
- Studies that explored diabetic care interventions conducted solely in the community setting without describing collaboration with health facilities will be excluded.

##### Types of Interventions

We will define the health system interventions using the modification from the framework by Witter et al(18) and a descriptive approach employed by Byinringiro et al(19)

- Studies would be eligible if the interventions address the following health system inputs (i.e., Service delivery, health workforce, supply chain, health information systems, financing governance) in the following ways;
- Service delivery. WHO defines service delivery as the activities that directly provide safe, effective, and high-quality health services to patients in need. Thus, we will consider the studies to enhance the capacity of this aspect of the health systems if the target of the intervention was to enhance patients access to health services for diabetes like screening, treatment, and follow-up, either through equitable distribution of care services, reduction of out-patient waiting time, revision of time allocated for services, or integrating the delivery of diabetes care with other established health services like HIV or TB.
- Health workforce. The interventions of interest in this aspect of the health systems would focus on healthcare providers who manage diabetes. The interventions include strategies to increase the number of providers, improve provider knowledge and implementation of diabetes management guidelines, address provider decision support systems, promote teamwork, and institute task sharing and/or task shifting strategies to include providers who do not normally perform certain diabetes management tasks like prescription of medications.
- Supply chain. In this aspect of the health systems, we will include studies with interventions that target enhancing the capacity of the procurement systems to ensure the availability of anti-diabetic medications, availability and maintenance of calibrated glucometers, availability of consumables to conduct screening and other diabetes investigations, patient follow-up technologies like use of short message systems, and treatment guidelines.
- Health information systems. We will consider interventions to enhance the capacity of health information systems if they studied an element of the process of patient data collection, analysis, sharing, and use to improve outcomes of interest. Interventions that explore the use of patient registries and information between patients and providers and among providers will be included
- Financing. The financing aspect of the health systems, as defined in the WHO building blocks, has the most significant relevance on the national macro-level health systems. It also plays an impact on the micro-level health systems’ service delivery and patients’ outcomes. We will consider studies where the interventions aimed at reducing patients’ out of pocket spending or funding of diabetes care at the health facilities through national and sub-national spending on health insurance premiums, or other relevant financial reliefs.
- Governance. This aspect has direct and indirect associations with the other five building blocks of the health systems. We will consider studies to address leadership and governance if the intervention promote a facility’s leadership awareness of the burden of poor diabetes management, or if the intervention uses strategic planning and implementation of national diabetes management protocols in a health care facility, explores the accountability measures at the health facility, applies regular performance appraisal and planning for improvement, integrates supportive mentorship to lower primary health facilities, institutes a patient feedback collection and response system, or examines the leadership allocation of funds for diabetes management. We will also consider interventions where leadership joins or collaborate with other health facilities or local/international/national/private/non-governmental organizations to manage diabetes.
- Only studies that piloted interventions with a minimum follow-up period of six months will be eligible.
- We will exclude studies that examined only patient-level or community-level lifestyle/behavioral changes or other non-conventional medical/non-medical interventions for diabetes management.

#### Comparison

Intervention groups would be compared with corresponding groups where there were no strategies to enhance any of the health systems inputs with respect to diabetes care.

#### Outcome

We will evaluate the impact of the interventions on any of the following outcomes:

- Good glycemic control. Defined by the American college of endocrinologists as glycated hemoglobin (HbA1c %) levels below 7% or fasting blood sugar levels below 110mg/dl (6.1 mmol/l)(4).
- Diabetes awareness. Defined as persons with clinically measured diabetes (either with HbA1c or fasting blood sugar levels), who have been diagnosed by a physician or health provider with appropriate training to make a diagnosis.
- Treatment Adherence. Defined as consistently complying with a treatment plan (either an oral anti-diabetic or insulin, or a life style plan) as prescribed/advised/implemented by a health care provider (22).
- Type 2 diabetes service availability and patient’s access to these services

#### Language

We will include articles published in English or French. These two are the main languages spoken in West Africa.

**S1 Table.** PICOS eligibility.

|  |  |
| --- | --- |
| **Population** | Adults aged 18 years or more in West Africa |
| **Intervention** | System-level interventions that targets the health system inputs in the Witter et al health system frame work within primary health facilities |
| **Comparison** | Corresponding groups where there were no strategies to enhance any of the health systems inputs with respect to diabetes care |
| **Outcome** | Main outcome is glycaemic control. Other health outcomes include: awareness, treatment adherence, type 2 diabetes services availability and patient access. |
| **Study design** | Randomized Controlled Trials (RCTs), Clinical Controlled trials (CCTs), quasi experiments |

### Search Strategy

#### Bibliographic databases

We will search the following databases PubMed, Google scholar and Cumulated Index to Nursing and Allied Health Literature (CINAHL) between January 2000 to June 2023 and Car.info from inception to June 2023. This time frame is selected to encompass studies that would have likely benefited from recent development and improvement in RCTs, CCTs and quasi-experimental studies. We will define the search terms for primary health care of diabetes at the facility level by borrowing and modifying the search strategy published in a systematic review of health systems interventions for hypertension by Byiringiro et al(19) These search terms will likely encompass the primary health care practices for diabetes management in our setting of interest.

The search strategy consists of intersections between the following medical subject headings (MeSH); (“Health Services,” Delivery of Health care,” “Primary Health Care,” “Health Facilities,” “Health Care Facilities, Manpower, and Services,” “Healthcare Financing,” “Insurance, Health, Reimbursement, “Health Information Systems,” “Equipment and Supplies”); and intersected with type 2 diabetes (“Diabetes mellitus, type 2,” “Glycemic Control,” “Diabetes Complications,” “Hyperglycemia,”) and West African countries (“African, Western”). The full search strategy is presented in S2 Table

#### Searching other resources

We will check the bibliographies of all relevant studies to identify other potential publications. The references of any relevant systematic reviews and scoping reviews identified will also be scrutinized for relevant trials.

We will also contact the authors of articles whose full text is not easily accessible after searching through multiple online databases and where there are questions related to the results of the study or trial design, we will seek confirmation on the information that we extract from their studies. In case of no feedback from the authors, the corresponding studies will be excluded. Also, studies that do not measure any of the outcome variables (glycaemic control, adherence, and awareness) will be excluded. Where there are duplicates for studies published in more than one paper, the most comprehensive one reporting the largest sample size will be considered.

### Study selection

Two reviewers will independently scan the titles and abstracts of all records yielded by the search to determine their eligibility for full-text screening and their full articles will be assessed through the databases and imported into the Rayyan screening software(22).

The full text of all potentially eligible articles will be screened and assessed for inclusion into the review, by a trained reviewer, using a pre-specified eligibility form based on the inclusion criteria (S3 Fig). At least one paper will be piloted before the main selection. Disagreements will be resolved after reaching a consensus or through discussion with a third reviewer. Neither of the review authors will be blind to the authors, institutions or journal titles of potential articles. We will present the number of included studies and the number of excluded studies using the PRISMA flow chart (S3 Fig)

### Data Extraction and management

Two reviewers will independently extract study characteristics and record them on pre-designed forms (S3 Table). The form would first be piloted on two included studies to ensure information is captured in a standard manner. We will extract data from each study on the first author, year of publication, language, study design, setting (country and facility level), health system domain and intervention explored, and outcomes. Disagreements will be resolved in consultation with the third reviewer.

### Dealing with missing Data

We will contact the authors of included studies for which data related to study methods, and outcomes are unclear or missing.

### Risk of Bias and quality Assessment of included studies

To assess for possible risk of bias in the included studies, we will collect information using the Cochrane Collaboration tool for assessing Risk of Bias(23)in a study (S4 Fig). The risk of bias tool would be implemented on each study and scored as high, low, or unclear risk (where there is insufficient detail reported), for the following domains: random sequence generation (selection bias); allocation concealment; blinding of participants and personnel (performance bias); blinding of outcome assessment (detection bias); incomplete outcome data (attrition bias); selective reporting (reporting bias). These scores would be assigned independently by the two review authors. Disagreements would be solved by discussion and consultation with a third reviewer. A table representing the risk of bias assessments within and across studies will be computed using Cochrane’s Review Manager (Version 5.3)

### Data synthesis and Analysis

We will conduct a narrative synthesis of the health system interventions identified from the studies by creating a table to compare the PICO elements. We will classify the outcomes into the following groups; glycemic control, diabetes awareness and treatment adherence. We will use relative risk to report the effect measures of the interventions on diabetes outcomes and compare them across all included studies. Our findings would be reported using tables generated from Excel.

### Assessment of confidence in cumulative estimate

The Grading of Recommendations, Assessment, Development, and Evaluation (GRADE)(24) approach will be implemented to assess the quality of the evidence for any of the outcomes. We will report a table on the GRADE evidence profile for the quality of the studies on the following domains: 1) risk of bias 2) inconsistency 3) indirectness 4) imprecision 5) publication bias 6) magnitude of effect 7) residual 8) dose-response 9) overall GRADE quality scored as very low (very uncertain about the estimate of effect), low (further research is very likely to change the estimate of effect, moderate (further research may likely change the estimate of effect), high quality (i.e., further research is very unlikely to change our confidence in the estimate of effect). (S5 Fig)

### Ethical considerations

Ethical approval will not be needed for this review protocol because data will be extracted from published studies and there will be no concerns about privacy.

## Discussion

By the end of this review, we aim to provide a report on integrated system-level interventions in the health systems of primary health facilities aimed at improving the delivery and quality of type 2 diabetes services as well as patients access to these services. We will further explore effectiveness of these interventions on the impact on patients’ glycemic control for at least 6 months. We would then outline areas of strengths and gaps in already existing interventions for the purposes of health policies, co-production of interventions and wide-spread uptake in the entire region.

### Strengths and Limitations

This review will provide valuable insights into the latest health system interventions for the management of type 2 diabetes in West Africa and explore the effect of integrated system-level interventions on provision of care for type 2 diabetes. The literature search would be conducted in only English and French thus we are likely to miss out on relevant studies in other languages. E.g., Portuguese.

### Dissemination plans

Results from this review would be disseminated through academic journals, policy briefs, stake holder and intervention co-production workshops.

### Dealing with Amendments

The corresponding author, Eugene Paa Kofi Bondzie, is responsible for any amendments and update of this review protocol.

## Authors’ contributions

**Study protocol conceptualization and design**: Eugene Paa Kofi Bondzie (EPKB), Yasmin Jahan (YJ), Dina Balabanova (DB).

**Supervision**: Irene Agyepong (IA), Edward Antwi (ED), Tolib Mirzoev (TM), Tony Danso-Appiah (TDA).

**Validation**: Dina Balabanova (DB), Irene Agyepong (IA), Tolib Mirzoev (TM).

**Original draft writing**: Eugene Paa Kofi Bondzie (EPKB)

**Review and editing**: Eugene Paa Kofi Bondzie (EPKB), Yasmin Jahan (EPKB)

## Data Availability

No datasets were generated or analysed during the current study. All relevant data from this study will be made available upon study completion.

## Acknowledgements

We would like to acknowledge Mary Pomaa Agyekum, the librarian who helped in the development of the search strategy.

## Supporting Information

S1 Checklist. Prisma-P 2015 checklist

(DOCX)

S2 Table. Search strategy for PubMed

(DOCX)

S2 Fig. Study selection flow chart

(DOCX)

S3 Fig. PRISMA flow chart

(DOCX)

S3 Table. Data extraction Form

(DOCX)

S4 Fig. Cochrane Risk of Bias Tool

(PDF)

S5 Fig. The Grading of Recommendations, Assessment, Development, and Evaluation (GRADE) Assessment Tool

(PDF)

